# Feasibility, acceptability and clinical outcomes of a real-world, regional lung cancer prehabilitation programme for patients undergoing curative intent radiotherapy

**DOI:** 10.1101/2025.01.11.25320256

**Authors:** Ewan Gourlay, Kathryn Banfill, Zoe Merchant, Patrick Goodley, Louise Brown, Jack Murphy, John Moore, Matthew Evison

## Abstract

**Introduction:** Prehabilitation improves both physiological measurements and clinical outcomes for patients undergoing surgery for lung cancer. The feasibility, acceptability and efficacy of prehabilitation for patients having curative-intent radiotherapy for lung cancer is uncertain.

**Methods:** Prehab4Cancer (P4C) is a regional, community-based prehabilitation service for patients with cancer in Greater Manchester in the United Kingdom. We present an evaluation of the P4C service for patients undergoing curative-intent radiotherapy for lung cancer over a 2-year period. Feasibility was evaluated against prespecified key performance indicators. Objective physiological and subjective functional assessments were recorded before and after completion. Effects on mortality in comparison to a non-prehab cohort were assessed in an exploratory regression analysis.

**Results:** A total of 381 patients were referred to P4C via a web-based portal. 86% (329/381) were contacted by phone and 73% (279/381) completed an initial assessment. 45% (172/381) completed the prehab programme with a median of 7 (IQR 3-11) sessions during a median time to commencing treatment of 23 days (IQR 16 – 34). Median time from referral to assessment was 4 days (IQR 3 – 6) and 86% (239/279) were completed within 7 days of referral. Six-minute Walk Tests improved by an average of 30m (95% CI 20.6–41m, p=<0.001). 16% (n = 21/132) of participants initially score “Medium” or “High” on IPAQ for weekly physical activity, improving to 52% (n = 66/132) on programme completion. Participants had a median reduction in score of −2.5 (95% CI −3.0 - −1.5) in WHODAS 2.0. There was an 8% reduction in 1-year mortality in patients completing prehab (11%, 17/160) vs those that did not complete prehab (19%, 30/158, OR 0.5, 95% CI 0.24–1.00, p=0.054 after adjustment for age, gender, performance status and cancer staging).

**Conclusions:** The P4C programme has demonstrated feasibility, acceptability and effectiveness in patients with lung cancer undergoing curative-intent radiotherapy.

## 1. Introduction

Lung cancer accounts for approximately one fifth of cancer-related mortality in the United Kingdom, with nearly 35,000 deaths per year (1). While surgical resection, when feasible, remains the intervention with best outcomes for long term survival, there are a large number of patients for whom this is not appropriate either due to anatomy or comorbidity (2). The use of radiotherapy as a curative-intent treatment modality has increased, especially the use of stereotactic ablative radiotherapy (SABR) for early stage tumours (3). Radiotherapy was further expanded during the COVID-19 pandemic period due to limitations in surgical capacity (4). Prehabilitation (prehab), in the context of cancer, refers to multimodal assessment and intervention in the time between diagnosis and definitive treatment, with the aim of improving both physical and psychological health (5). There are multiple studies demonstrating the benefit of prehab for patients undergoing lung cancer surgery, with improvements in functional capacity and reductions in post-operative complications (6)(7). In contrast to surgical intervention, the evidence base for prehab for patients undergoing curative-intent radiotherapy treatment for lung cancer is limited. Furthermore, the National Lung Cancer Getting It Right First Time (GIRFT) Specialty report recommended ‘*Access to prehabilitation should be available for all patient undergoing radical intent treatment but we have found this varies significantly*’(8). There is, therefore, an urgent need to develop the evidence-base and understand the real-world feasibility and acceptability of delivering prehabilitation in this patient cohort.

The Greater Manchester (GM) Prehab4Cancer (P4C) programme is a regional service offering individualised prehab and rehabilitation (rehab) programmes for patients undergoing surgery for lung, colorectal and oesophagogastric cancers. P4C has demonstrated feasibility at scale, as well as significant improvements in both subjective functional and objective physiological measures for lung cancer patients treated with surgery (9). The P4C service was expanded to include lung cancer patients treated with curative-intent radiotherapy in 2020. The outcomes of this expansion are analysed here.

## 2. Methods

### 2.1 Service Background

The GM ceremonial county and combined authority is located in the North-West of England, with a population of ∼2.8million. There are approximately 2500 lung cancer cases diagnosed each year within the region. Patients diagnosed with cancer in GM are on average less active and have poorer health compared to other UK areas (10). P4C is organised in collaboration between the ‘GM Cancer’ alliance which leads cancer care across 11 acute hospitals and ‘GM Active’, a collaboration of 12 publicly owned leisure and community organisations across the city-region (11). As an expansion of the Enhanced Recovery After Surgery Plus (ERAS+) programme, the service was launched originally for patients treated with surgery for lung cancer in 2019, with subsequent expansion of eligibility to patients having radiotherapy in 2020. A more detailed description of the inception of P4C were previously covered in detail by *Moore* (12). The P4C team during this study period consisted of one program manager, six exercise specialists, three qualified exercise instructors and a referral co-ordinator.

### 2.2 Pathway & Intervention

Patients were referred to P4C at the point of MDT discussion utilising a single access online portal. Eligibility criteria for this pathway is: aged 18 or older; MDT agreed diagnosis of primary lung cancer eligible for curative intent treatment including radiotherapy; ECOG Performance Status (PS) ≤2; Rockwood Clinical Frailty Score (CFS) ≤5; and suitable for a community prehab and recovery programme in the opinion of the referring clinician. Referred patients are contacted initially via telephone to offer an initial face-to-face assessment at one of 17 assessment clinics across GM. Alongside a medical history, baseline assessments are taken for physiological measures including Incremental Shuttle Walk Test or 6 Minute Walk test, 60 second Sit-to-Stand, Hand-Grip Dynamometry and Body Mass Index. Subjective functional assessments are performed using 12 item WHO Disability Schedule 2.0 (WHODAS), Self-Efficacy for Exercise (SEE), International Physical Exercise Questionnaire (IPAQ) and European Quality of Life Five Dimensions instrument (EQ-5D-5L). Following initial assessment, an individualised prehab programme is prescribed. A repeat assessment is performed prior to starting oncological therapy. Patients are offered continued intervention in the period while receiving radiotherapy and if received, chemotherapy. These exercise prescriptions are at a reduced intensity and with careful monitoring to not negatively impact oncological treatment. Patients are also offered a 12-week recovery service after treatment. The individual exercise prescriptions in the prehabilitation period generated for each patient encompassing aerobic and resistance training. Patients are placed on a “Targeted” or “Universal” pathway depending upon their baseline fitness and the perceived need for supervision to support exercise intensity or other aspects of prehab. Patients stratified to the “Targeted” pathway receive three supervised sessions per week generally delivered as small group sessions across the GM Active community leisure facilities. Patients stratified to the “Universal” pathway generally undertake unsupervised prescribed exercise with weekly review by the exercise specialist, however they can also join group supervised exercise sessions. Both programmes provide intensity escalation as the patient improves fitness through prehab. Nutrition and psychological wellbeing are considered at all assessment points of service delivery (baseline, prehab completion, post treatment/pre-recovery phase and post-recovery/discharge). The Patient Global Subjective Assessment Scale (PG-SGA) is used to screen nutritional risk. Low risk patients (0-1 PG-SGA score) receive a P4C diet plan devised by the GM P4C nutritional subgroup (including specialist dietitians and other members of the MDT), moderate risk patients (2-3 PG-SGA score) are provided with the more comprehensive ‘Eating Help Yourself’ booklet (designed by the Christie NHS hospital) and those screened to be high risk of malnutrition (4 and above PG-SGA score) are escalated back to the referrer for urgent dietetic assessment. The wellbeing offer includes peer support from fellow patients accessing the service, psycho-social support from P4C team members and the evidence based psychological benefits of engaging in regular exercise, including improvements in mood disturbance and reduction of anxiety (13)(14), both common sequelae of being diagnosed with cancer (15). The P4C exercise specialists engage in monthly reflective sessions with a cancer-specialist consultant clinical psychologist to support their effective delivery of the wellbeing modality within the P4C service. Finally, patients deemed to have high mental health need(s) are signposted to primary care local services, with concerns raised communicated back to their named referrer. During the second wave of the COVID-19 pandemic, modifications were made to minimise risk of infection. Remote assessments were performed and amended exercise prescriptions were made to account for the closure of leisure centres. Following the relaxing of COVID-19 restrictions in July 2021, session in leisure centres were restarted, though the options of home exercise or blended (i.e. Home and Gym) were also available. This evaluation covered referrals during a 2-year period from 1^st^ October 2020 until 30^th^ September 2022.

### 2.3 Methodology

The P4C programme is underpinned by a service database that records all referrals, demographic & clinical information, the outcome of initial telephone contact & initial assessment, baseline physiological and functional assessments, all individual prehab-rehab sessions and results of any repeat physiological & functional assessments. This service data was linked to clinical outcomes via the GM tertiary oncology centre patient records to provide treatment and cancer-related outcomes such as toxicity and survival. Key Performance Indicators (KPIs), used as markers of feasibility, uptake and acceptability, initially described by *Bradley et al* for assessment of the surgical resection lung cancer cohort have been adapted for use here (8). In a sub-group analysis, patients that completed curative-intent radiotherapy and completed the full course of prehabilitation prior to treatment were compared to patients that completed curative-intent radiotherapy that did not complete prehab for toxicity and survival outcomes. All relevant patients referred to P4C were included in initial KPI analysis. Patients whose cancer was not compatible with radical therapy prior to starting oncological therapy (i.e. M1 disease), or who did not start therapy (e.g. decision for active surveillance or revised diagnosis) were excluded from the analysis of physiological and functional assessments. Analysis of toxicity and mortality also excluded those who were medically assessed as ineligible from the P4C telephone or first clinic assessment to avoid biasing the ‘did not complete’ P4C group with increased morbidity.

### 2.4 Statistical analysis

For statistical testing, paired and un-paired T-tests were used for continuous parametric data, Wilcoxon Signed-rank for non-parametric data and Pearsons χ2 for categorical data. Analysis was performed using Graphpad Prism Version 9.5.1 (528), January 24, 2023. Multivariable regression analysis was performed using R Studio (Version 2023.09.1+494). EQ-5D-5L index scores were calculated using the England specific 2018 value set (16).

### 2.5 Patient & public involvement

Two GM patient representatives were members of the GM Prehab4cancer Steering Group with additional patient representatives in the Prehab4cancer Lung Sub-group. These patient representatives co-designed the service delivery models and led the development of patient information materials and communications.

### 2.6 Ethical Approval

Formal ethical review was not required given the observational nature of this service evaluation, confirmed through the Health Research Authority toolkit and at local review board.

## 3. Results

### 3.1 Patient characteristics & Uptake

During the prescribed analysis period 381 patients expected to undergo curative-intent radiotherapy were referred to the P4C programme. The average age was 74 (95% CI 68-78), 52% (176/381) were male, 60% (228/381) were stage I/II and 55% (208/381) were PS 0-1 (Table 1). Referrals to P4C were received from all 13 eligible hospital sites. In the first year, 149 patients were referred to P4C, rising to 232 patients in the second year. 14% (52/381) patients were unable to be contacted by telephone (Figure 1). Of the 329 patients initially contacted by telephone, 85% (279/329) completed an initial assessment. By intention to treat (ITT) analysis, 73% (279/381) completed first assessment. Median time from referral to initial assessment was 4 days (IQR 3 – 6). Assessment of outcomes against prespecified KPIs is illustrated in Table 2. Of the patients who underwent first assessment, excluding failed assessments, 64% (169/264) were stratified to the “Targeted” pathway and 33% (88/264) to “Universal” pathway. A total of 7% (27/381) of patients were deemed medically ineligible; 12 at initial phone contact and a further 15 at initial assessment. 172 patients completed the full P4C prehabilitation programme up until treatment commencing, equating to 45% (172/381) of all patients initially referred to the programme. Time from first assessment to initiation of oncological treatment was median 23 days (IQR 16 – 34). Patients who completed the P4C prehab phase undertook a median of 7 (IQR 3 – 11) individual exercise sessions. Ninety patients continued with the service after oncological therapy into the rehabilitation phase, of whom 76% (n = 69/90) completed a formal rehabilitation programme and assessment. For those who undertook both assessments, median duration was 98 days (IQR 88 – 118) and included a median 13 (IQR 4 – 24) prescribed exercise sessions. This cohort contained 7 patients who did not complete the prehab phase due to proximity to treatment. For the sub-group analysis, a cohort of 160 patients that completed curative-intent radiotherapy and completed prehabilitation was compared to a cohort of 180 patients that completed curative-intent radiotherapy but did not complete prehab. The two cohorts were matched for age, gender, performance status, lung function and cancer staging (Table 1 & Supplementary Table 2).

**Figure 1.**
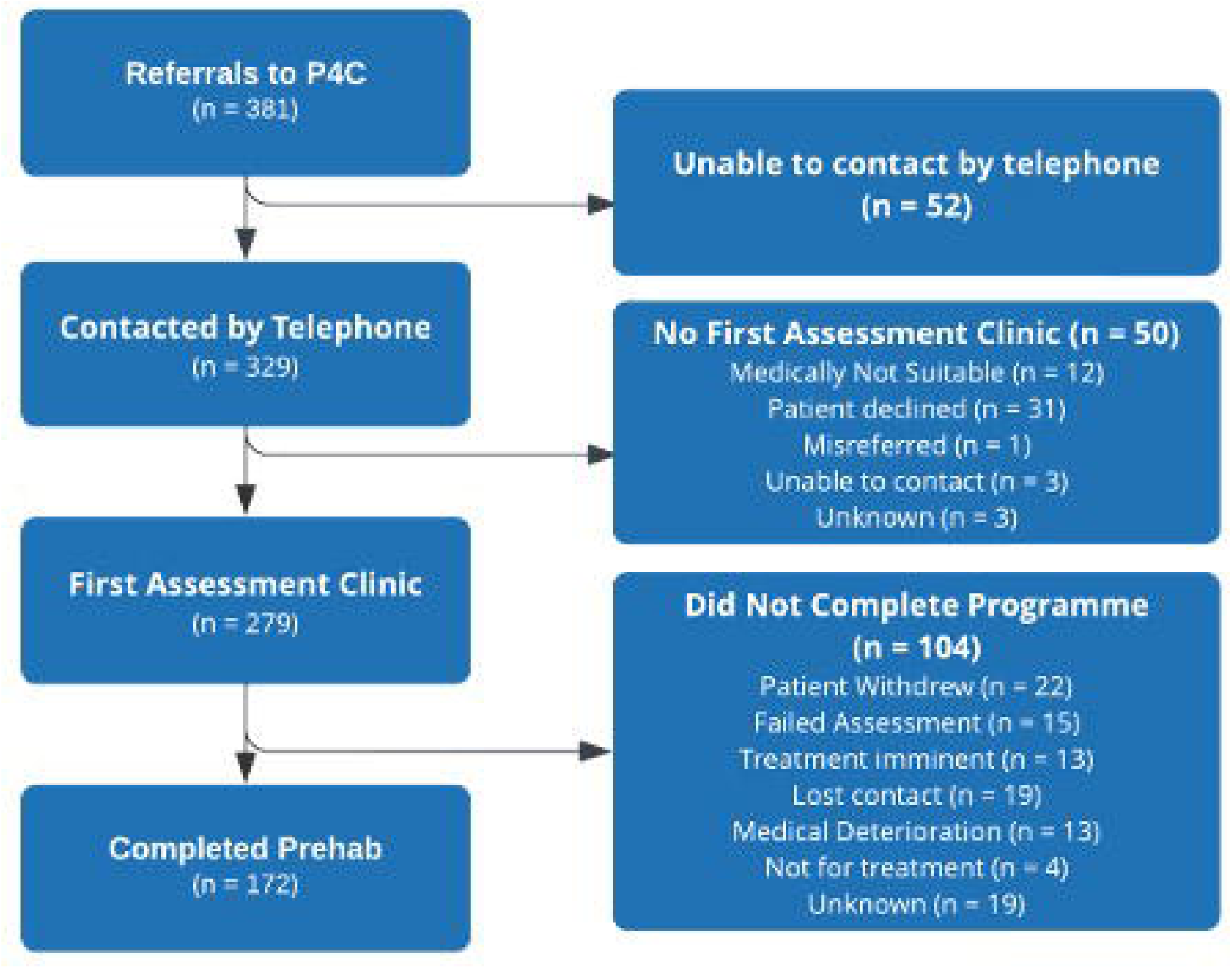
Patient progression through P4C pathway.

**Table 1.** Patient characteristics.

| Variable | N | Overall, N = 340<sup>1</sup> | Completed P4C |  |
| --- | --- | --- | --- | --- |
|  |  |  | No, N = 180<sup>1</sup> | Yes, N = 160<sup>1</sup> |
| Age | 340 | 74 (68, 78) | 73 (68, 78) | 74 (68, 78) |
| Gender | 340 |  |  |  |
| Female |  | 164 (48%) | 82 (46%) | 82 (51%) |
| Male |  | 176 (52%) | 98 (54%) | 78 (49%) |
| Tumour Stage | 340 |  |  |  |
| I |  | 171 (50%) | 92 (51%) | 79 (49%) |
| II |  | 46 (14%) | 21 (12%) | 25 (16%) |
| III |  | 100 (29%) | 56 (31%) | 44 (28%) |
| Nodal Only |  | 12 (3.5%) | 8 (4.4%) | 4 (2.5%) |
| Synchronous Primaries |  | 11 (3.2%) | 3 (1.7%) | 8 (5.0%) |
| ECOG PS | 340 |  |  |  |
| 0 |  | 46 (14%) | 20 (11%) | 26 (16%) |
| 1 |  | 144 (42%) | 74 (41%) | 70 (44%) |
| 2 |  | 123 (36%) | 69 (38%) | 54 (34%) |
| 3 |  | 27 (7.9%) | 17 (9.4%) | 10 (6.3%) |
| FEV1 % Predicted | 330 | 71 (58, 86) | 70 (58, 86) | 72 (57, 86) |
| DLCO % Predicted | 316 | 56 (44, 69) | 55 (44, 68) | 57 (46, 70) |
| <sup>1</sup> Median (IQR); n (%) |  |  |  |  |

**Table 2:** Key Performance Indicators.

| KPI | Outcome Assessed | Red | Amber | Green | Outcome % (n) |
| --- | --- | --- | --- | --- | --- |
| 1 | Proportion of referrals successfully contacted by telephone (%) | <75 | 75-90 | >90 | 86 (329/381) |
| 2 | Proportion of patients completing a first assessment consultation after a successful telephone contact (%) | <50 | 50-75 | >75 | 85 (279/329) |
| 3 | Proportion of patients completing a first assessment consultation (ITT) (%) | <30 | 30-65 | >65 | 73 (279/381) |
| 4 | Proportion of first assessment clinics completed within 7 days of referral (%) | <50 | 50-75 | >75 | 86 (239/279) |
| 5 | Proportion of patients deemed medically unsuitable for P4C at first assessment clinic (%) | >20 | 10-20 | <10 | 5 (15/279) |
| 6 | Proportion of patients completing the P4C prehab phase after first assessment (%) | <50 | 50-75 | >75 | 61 (172/279) |
| 7 | Proportion of patients completing the P4C prehab phase (ITT) (%) | <25 | 25-50 | >50 | 45 (172/381) |

### 3.2 Oncological Therapy

Ninety-one percent (340/374) of patients referred to P4C started curative-intent treatment. Treatment data is unavailable for 7 patients. Fourteen received no treatment and 18 started treatments with palliative intent. (Supplementary Table 1). A total of 339 patients received curative intent radiotherapy, with 41% (139/339) treated with SABR and 21% (n = 70/339) treated with chemoradiotherapy (43 with concurrent chemoradiotherapy and 27 with sequential chemoradiotherapy). Full details of histology and radiotherapy fractionation are detailed in Supplementary Table 2.

### 3.3. Physiological & Functional Assessments

Statistically significant improvements were seen in both 6-minute walk tests (6MWT) (Mean difference +30m 95% CI 20.6–41m) and 60 second sit to stand (STS) (mean difference +3.3 95% CI 2.7–3.9, Figure 2, Table 3) comparing before and after prehab. A reduced number of paired physiological measures were assessed, in part due to remote assessments during COVID-19. Comparing initial physiological assessments for patients completing P4C vs those who did not, revealed no significant difference for 6MWT (completed P4C n = 23, did not complete P4C n = 38, p= 0.24) and STS (completed P4C n = 48, did not complete P4C n = 46, p = 0.25). A total of 6 paired HGD measures were available so were not further analysed.

**Figure 2.**
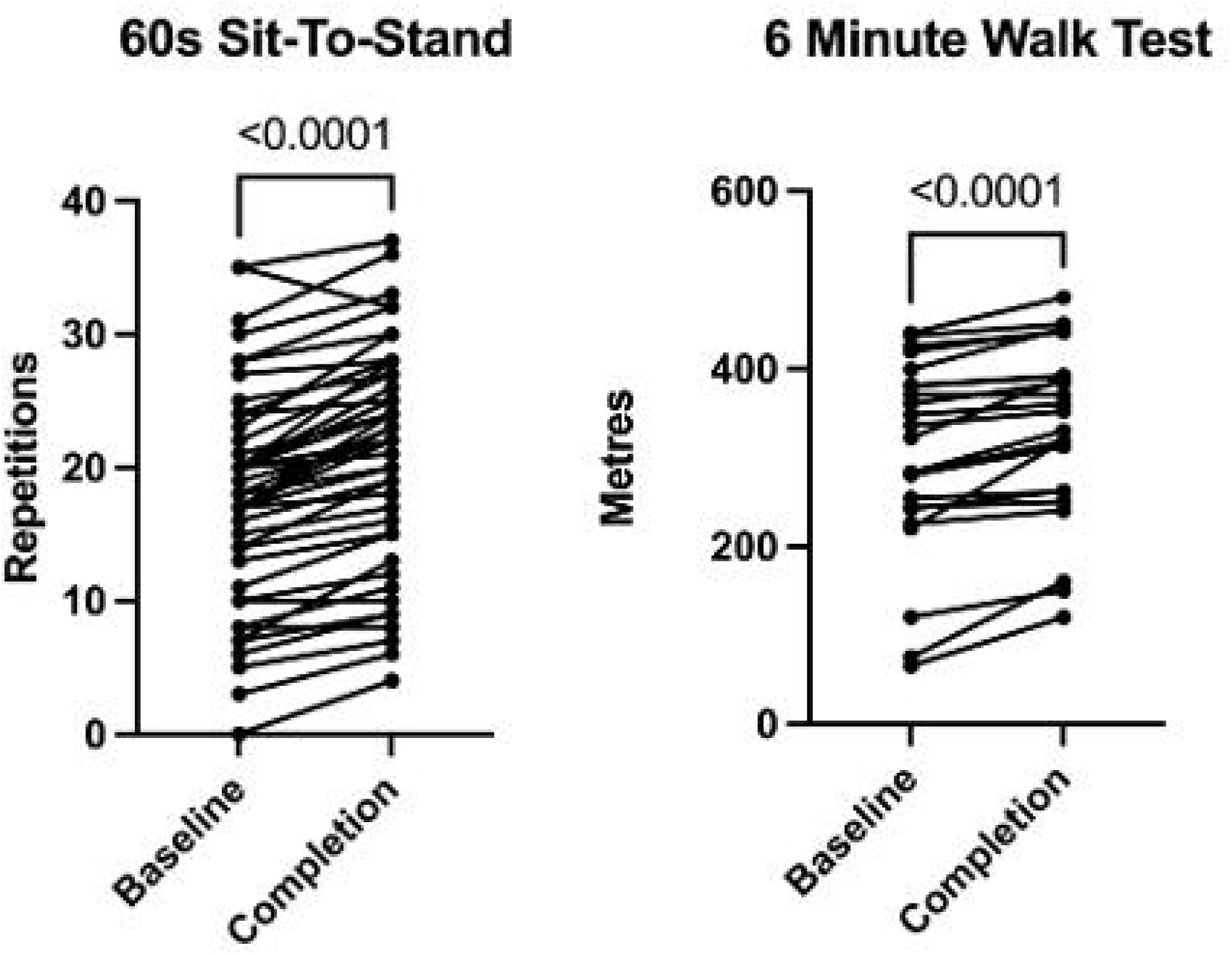
Objective Physiological paired measures.

**Table 3.** Physiological & Functional Assessments.

| Variable<sup>α</sup> | n | Initial Assessment (All)<sup>β</sup> | Initial Assessment (Paired only) | End of Prehab<sup>β</sup> | N paired results | Difference (95% CI)<sup>δ</sup> | P-value<sup>δ</sup> |
| --- | --- | --- | --- | --- | --- | --- | --- |
| 6MWT (m) | 72 | 277<br>(117) | 302<br>(106) | 333<br>(97) | 25 | 30.8<br>(20.6 – 41.0) | < 0.001 |
| STS (reps in 60s) | 104 | 16.6<br>(8.0) | 17.5<br>(8.1) | 20.7<br>(8.21) | 51 | 3.3<br>(2.7 – 3.9) | < 0.001 |
| WHODAS | 279 | 6<br>(2.5–14) | 6<br>(2–14) | 5.5<br>(2–1.8) | 132 | -2.5<br>(-3.0 - -1.5) | < 0.001 |
| SEE | 279 | 60<br>(45–72) | 58<br>(45–70) | 68<br>(57–78) | 129 | 11<br>(9.0 – 14.0) | < 0.001 |
| IPAQ, n (%) | 279 |  |  |  | 132 |  | < 0.001 |
| Low |  | 234<br>(84) | 111<br>(84) | 66<br>(48) |  |  |  |
| Medium |  | 40<br>(14) | 20<br>(15) | 53<br>(42) |  |  |  |
| High |  | 5<br>(2) | 1<br>(1) | 13<br>(10) |  |  |  |
| EQ-5D-5L | 279 | 0.796 (0.660 -0.922) | 0.796 (0.672 – 0.879) | 0.808 (0.701 – 0.922) | 132 | 0.041 (0.025 – 0.060) | < 0.001 |

α 6MWT, 6 minute walk test; STS, Sit-to-Stand test; WHODAS, WHO Disability Assessment Schedule 2.0; SEE, Self-Efficacy for Exercise Scale; IPAQ, International Physical Activity Questionnaire; EQ-5D-5L, EuroQol-5 Dimension-5 Level assessment;.
β Mean (standard deviation); Median (Interquartile Range)
δ Paired values assessed, Paired T-Test (parametric data), Wilcoxon Signed Rank (non-parametric data) Pearson's $\chi^2$ -squared (Categorical data)

**Table 4.** Toxicity & Mortality outcomes in prehab vs no prehab cohorts.

|  | Overall<br>N=318 | Completed Prehab<br>N= 160 | Did not complete<br>Prehab N = 158 |
| --- | --- | --- | --- |
| G2+ Toxicity | 155 (49%) | 75 (47%) | 80 (51%) |
| 90-day mortality | 12 (4%) | 5 (3%) | 7 (4%) |
| 365-day mortality | 47 (15%) | 17 (11%) | 30 (19%) |

Statistically significant improvements were seen in all subjective functional assessments performed when comparing paired results (Figure 3, Table 3). Comparing paired values, 16% (n = 21/132) of participants initially score “Medium” or “High” on IPAQ for weekly physical activity suggesting at least 3 high intensity or 5 moderate intensity exercise sessions per week. This proportion improved to 52% (n = 66/132) on completing the programme. Assessing disability using WHODAS 2.0, participants had a median reduction in score of −2.5 (95% CI −3.0 - −1.5), where 0 is no disability and 48 is maximum in all domains. The proportion of patients with WHODAS scores of ≥10, equivilent to top 10% of overal population for disability (17), reduced from 39% (n = 52/132) to 27% (n=36/132).

**Figure 3.**
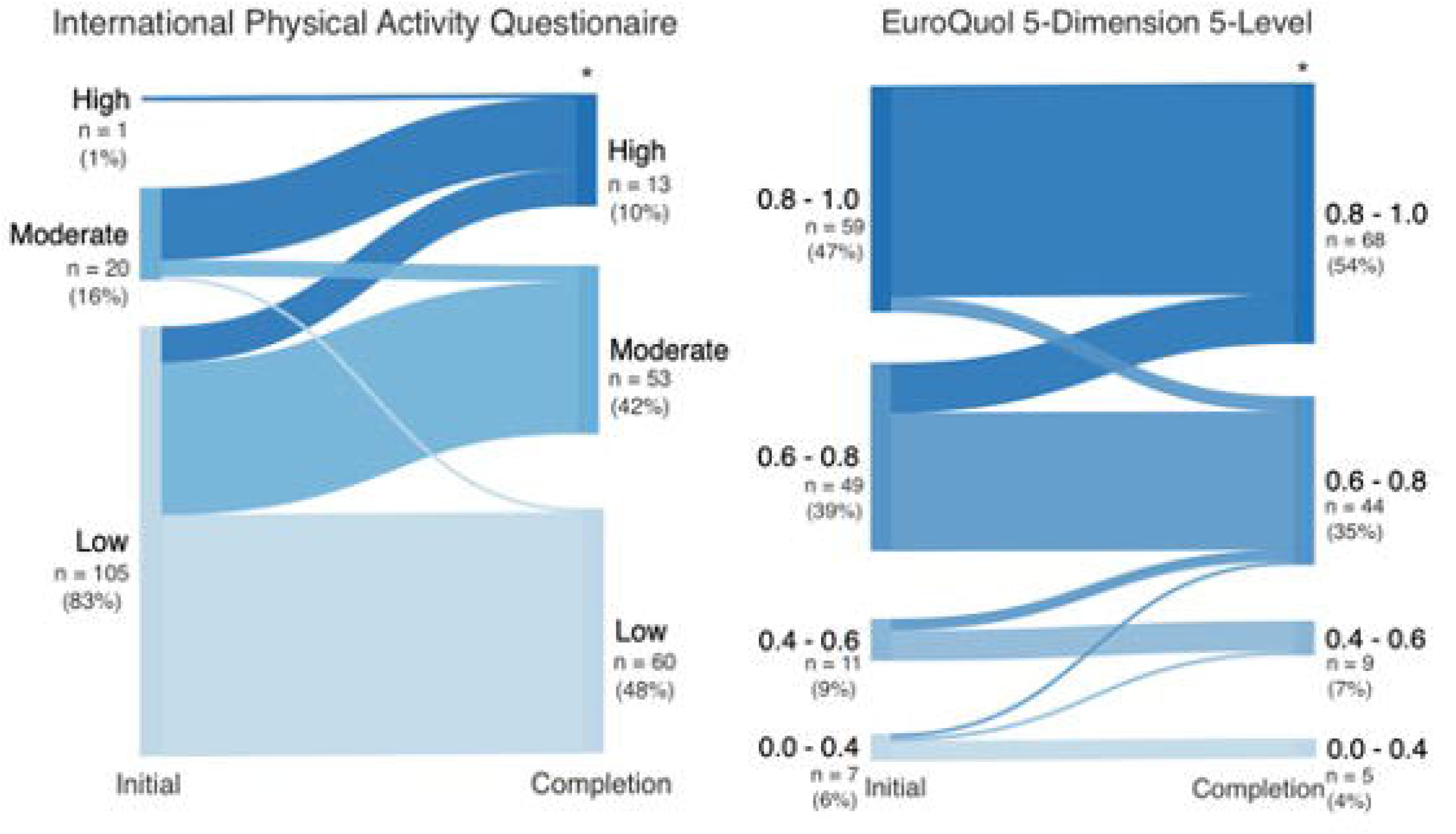
Selected Subjective Functional Assessments.

### 3.4 Universal vs targeted pathways

Participants allocated to the “Targeted” arm had significantly worse baseline physiological measurements (6MWT, STS) and subjective functional assessments (WHODAS, SEE, EQ-5D-5L) vs “Universal”. Those in the “Targeted” arm had a significantly greater (−2 (0-−2), p=< 0.001) change vs “Universal” in WHODAS assessment after completing P4C. There was no significant difference in other subjective functional assessments.

### 3.5 Toxicity & Mortality data

Mortality & toxicity analysis excluded 22 patients who started radical treatment but were assessed as medically unsuitable for P4C. In total 49% (n = 155/318) patients had a documented CTCAE grade 2 or higher toxicity documented during or in the 6 weeks following treatment. The most common being oesophagitis (n = 77), fatigue (n= 31), dyspnoea (n=17) and lung infection (n = 13). 90-day mortality was 4% (n = 12/318) while 365-day mortality was 15% (n= 47/318) overall (Table 5). Logistic regression models were constructed for toxicity and mortality. Completion of the P4C prehab programme had low univariable predictive value for grade 2 or higher toxicity (OR 0.86 95% CI (0.55 – 1.34) p = 0.50) and 90-day mortality (OR 0.70 (0.20 – 2.33) p = 0.54). Completion of P4C had an estimated OR of 0.50 (0.24 – 1.00) p=0.054 for 365-day mortality after adjustment for age, gender, performance status and cancer staging (Supplementary Tables 5-7). Fourteen patients had delays in radiotherapy course due to acute complications, of whom six completed the P4C programme.

### 3.6 COVID-19 impact on services

Analysis of patients referred to P4C before or after July 19^th^ 2021, the end of all UK COVID-19 restrictions, shows no difference between groups for subjective functional assessments (Supplementary Table 4). 60s Sit-to-Stand showed a significant improvement of 1.51 (95% CI 0.13 – 2.86) for the group seen after restrictions ended. Only 1 6MWT paired assessment occurred during the covid period so was not analysed.

## 4. Discussion

### 4.1 Summary of Key findings

The majority of patients referred to prehab4cancer in this cohort were contacted by phone (86%) and nearly three-quarters of referred patients completed a first assessment (73%). This initial assessment was completed in a median of 4 days and 86% were within 7 days. The proportion of referred patients that were ultimately deemed medical ineligible was low at 7%. Overall, 45% of referred patients completed prehab up to the point of treatment commencing, achieving a median of 7 prehab sessions in a median time of 23 days from initial assessment to treatment commencing. The 6MWT improved by an average of 30m. The proportion of participants undertaking at least 3 high intensity or 5 moderate intensity exercise sessions per week increased from 16% to 52% and the proportion of patients classified as within the 10% most disabled in the general population reduced from 39% to 27%. Completion of P4C had an estimated OR of 0.50 (0.24–1.00, p=0.054) for 365-day mortality after adjustment for age, gender, performance status and cancer staging.

### 4.2 Discussion

The first objective of this evaluation was to understand the feasibility and acceptability of prehabilitation in patients with lung cancer planned for curative-intent radiotherapy. This is especially important as patients on this treatment pathway are often older, frailer and have more co-morbidity. This could impact on the ability to complete physical activity as well as attend community sessions on a regular basis. Radiotherapy pathways are also faster than surgical pathways which has led some to question the feasibility of prehabilitation delivery within radiotherapy treatment pathways. Our evaluation has demonstrated a high-volume referral rate across a regional footprint (n=381 over a two-year period). An estimated 600 patients underwent radical radiotherapy for lung cancer in GM between Oct 2021–Sept 2022. In this timeframe there were 232 referrals for prehabilitation. It is not possible to truly understand the overall eligible population for prehabilitation given our inclusion criteria of PS ≤2, CFS ≤5 and deemed suitable for a community-based prehab programme. We can conclude however that these simple referral criteria do not lead to excessive referrals of medically ineligible patients that cannot complete community prehabilitation in this cohort (7% deemed ineligible) demonstrating a feasible service delivery model. The acceptability is high with 73% completing initial assessments, with these initial appointments happening within a median of 4 days (86% within 7 days) which facilitates an average of 7 sessions and 45% completing the full prehabilitation programme in the median pathway time of referral to treatment of 23 days. This compares favourably with the previously published surgical cohort undergoing surgery in GM with 74% completing initial assessments, with these initial appointments happening within a median of 8 days (48% within 7 days) which facilitates an average of 6 sessions and 48% completing the full prehabilitation programme in the median pathway time of referral to treatment of 36 days. Therefore, despite increasing age, frailty, comorbidity and faster treatment pathways for radiotherapy, this evaluation has demonstrated it is feasible to deliver rapid access to prehabilitation and it is acceptable at scale.

As expected, the ‘Targeted’ group had significantly worse physiological and subjective functional baseline assessments. Those in the ‘Targeted’ group had a significant improvement in the WHODAS assessment, but not the other subjective assessments. On review of the individual data, this difference appears to be driven by a subgroup of the ‘Targeted’ cohort of patients who had high levels of initial disability and significant improvement on completing P4C. This demonstrates that prehab can have the greatest effect for the least fit patients, of whom there are more in the radiotherapy cohort (i.e. having SABR because not fit for surgery). This agrees with data from surgically treated cohorts showing greatest relative improvements in outcomes following prehab in those with initially poorer health and fitness.

During the COVID-19 pandemic, in view of reducing transmission and due to legal requirements, face to face assessments and exercise sessions were not possible. While some tests such as the Sit to Stand test have been demonstrated to be reliably assessed remotely in older adults(18), others such as the 6MWT/ISWT or hand grip dynametry were not possible and as such reduced data is available during this time for analysis. Furthermore, all exercise had to be performed in the home rather than in gym leisure centres. When comparing those who completed P4C during and after Covid-19 restrictions, there was no significant difference in change for subjective functional assessments but there was a greater improvement in Sit-To-Stand test for those participating after restrictions finished. This could suggest that in person sessions are more beneficial for specifically improving cardiovascular or muscular conditioning, but remote interventions are as effective for holistic improvement.

Analysis of toxicity did not reveal a significant difference between groups completing P4C vs those who did not. Toxicity was assessed using CTCAE v5 (19), with the most common toxicity documented being oesophagitis. Oesophagitis, if symptomatic and thus documented by a clinician would be grade 2 or higher, by definition. After adjustment for confounding variables, 365-day mortality showed a trend towards significance, with benefit for those patients undergoing prehab. In other respiratory conditions such as COPD, participation in pulmonary rehabilitation programmes, which are similar to the P4C programme, have been shown to be associated with a significant benefit(20)(21). It may be that general improvements in health and wellbeing achieved via completing P4C offers a survival benefit for non-cancer mortality. Though there is a risk of bias contributing to this effect, this signal warrants further future investigation.

### 4.3 Context within published literature

The published literature on prehabilitation for patients with lung cancer lung cancer undergoing curative-intent radiotherapy is limited. *Egegaard et al* reported a feasibility study of a daily exercise programme in patients with lung cancer. Fifteen patients were randomised to daily exercise versus usual care with eight patients randomised to the intervention arm. In these eight patients, the 6MWT improved by an average of 33m (similar to our finding of an average 30m improvement) and there were improvements in treatment-related fatigue(22). *Goldsmith et al* reported a prospective study of prehabilitation in patients with lung cancer being worked up for curative-intent treatment. The study cohort was 216 patients and within this there 26 patients treated with radiotherapy and the specific outcomes for this cohort were not provided(23). Overall, therefore, there is a total of 34 patients in the published literature for prehabilitation in patients with lung cancer undergoing curative intent radiotherapy. Our study of 381 patients referred for prehabilitation prior to curative-intent radiotherapy, therefore, provides critically important data to the published literature to understand feasibility, acceptability and clinical outcomes in this patient cohort. To our knowledge, this programme represents the first prehab service for patients undergoing curative intent radiotherapy operated at scale across a region and cancer alliance.

### 4.4 Limitations

There are inherent limitations of service evaluations versus prospective randomised controlled trials. There is likely to be selection bias inherent to those completing the programme despite there being no statistically significant differences across several clinical and demographic factors assessed (Table 1 & Supplementary Table 2). One single centre RCT of prehab in lung cancer patients treated with radiotherapy “PREHABS” is currently in progress (24), which will add to understanding of the impact of prehab in this cohort. Though statistically significant improvements were seen in all assessments with enough measures to analyse, some changes were small in real terms. There is no validated data available to set thresholds for minimum clinically important differences (MCID) in appropriate cohorts (e.g. patients with lung cancer) with the exception of the 6MWT which was on average achieved in this cohort (25).

## 5. Conclusion

P4C is the first real-world prehab programme to include a curative-intent radiotherapy treated lung cancer cohort across a cancer alliance. It has demonstrated feasibility and acceptability at a service level with its expansion from surgical patients to the new cohort of patients having radiotherapy. Patients completing the programme showed significant improvements in objective physiological and subjective functional assessments, including via remote delivery during COVID-19 restrictions. Those patients with worst function prior to starting prehab had the largest improvement in health and disability, demonstrating the importance of prehab in less fit patients who have radiotherapy rather than surgery.

## CREDIT Authorship statement

**Ewan Gourlay:** Writing-Original draft preparation, Writing – reviewing and editing, Visualisation, Data Curation, Formal Analysis, **Kathryn Banfill**: Methodology, Writing – reviewing and editing, **Zoe Merchant:** Methodology, Investigation, Writing – reviewing and editing **Patrick Goodley:** Formal Analysis, Writing – reviewing and editing, **Louise Brown:** Writing – reviewing and editing, **Jack Murphy:** Data Curation, Writing – reviewing and editing **John Moore:** Conceptualisation, Writing – reviewing and editing, Supervision, **Matthew Evison:** Conceptualisation, Methodology, Writing – reviewing and editing, Supervision, Funding acquisition.

## Data Availability

All data produced in the present study are available upon reasonable request to the authors

## Acknowledgements

We would like to acknowledge and thank GM Cancer, GM Active, the Prehab4Cancer team and members of the GM lung pathway board.

