## Supplementary Tables 1-8 for "Feasibility, acceptability and clinical outcomes of a real-world, regional lung cancer prehabilitation programme for patients undergoing curative intent radiotherapy"

Supplementary Material

**Supplementary Table 1** – Reason for exclusion from subsequent analysis

| Exclusion Reason | | |
| --- | --- | --- |
| **Category** | **Reason** | **N (%)** |
| Administrative | Incorrect NHS Number | 6 (15%) |
| Administrative | Out of region treatment | 1 (3%) |
| Not Treated | Active Surveillance | 7 (18%) |
| Not Treated | Died before treatment | 1 (3%) |
| Not Treated | Patient declined treatment | 1 (3%) |
| Not Treated | Non cancer health | 3 (8%) |
| Not Treated | Cancer diagnosis revised | 2 (5%) |
| Palliative Treatment | Upstaged prior to treatment | 6 (17%) |
| Palliative Treatment | General health | 3 (5.1%) |
| Palliative Treatment | Metastatic at referral | 9 (23%) |
| Total |  | 39 |

**Supplementary Table 2. Lung Cancer histology & treatment modality**

| **Variable** | **N** | **Overall**, N = 340*^1^* | **Completed P4C** | |
| --- | --- | --- | --- | --- |
|  |  |  | **No**, N = 180*^1^* | **Yes**, N = 160*^1^* |
| Histology | 340 |  |  |  |
| Adenocarcinoma |  | 99 (29%) | 50 (28%) | 49 (31%) |
| NSCLC_NOS |  | 24 (7.1%) | 9 (5.0%) | 15 (9.4%) |
| Other |  | 3 (0.9%) | 0 (0%) | 3 (1.9%) |
| Radiological Diagnosis |  | 92 (27%) | 50 (28%) | 42 (26%) |
| Small Cell |  | 14 (4.1%) | 9 (5.0%) | 5 (3.1%) |
| Squamous |  | 105 (31%) | 61 (34%) | 44 (28%) |
| Synchronous Primaries |  | 3 (0.9%) | 1 (0.6%) | 2 (1.3%) |
| Radiotherapy Fractionation | 339 |  |  |  |
| ≤8 # (SABR) |  | 139 (41%) | 70 (39%) | 69 (43%) |
| 50-60 Gy in 15-20 # |  | 150 (44%) | 87 (49%) | 63 (39%) |
| 60-66 Gy in 30-33 # |  | 41 (12%) | 17 (9.5%) | 24 (15%) |
| Other |  | 9 (2.7%) | 5 (2.8%) | 4 (2.5%) |
| SACT | 70 |  |  |  |
| Concurrent |  | 43 (61%) | 18 (49%) | 25 (76%) |
| Sequential |  | 27 (39%) | 19 (51%) | 8 (24%) |
| *^1^* n (%) | | | | |

**Supplementary Table 3.** Initial assessment by allocated prehab arms

| Variable^α^ | Universal | | Treated | |  |  |
| --- | --- | --- | --- | --- | --- | --- |
|  | n | Average | n | Average | Difference  (95% CI) | P-value |
| STS (60s) | 27 | 19.9 (7.1) | 44 | 15.9 (8.2) | - 4.06 (-7.882 - -0.242) | 0.038 |
| 6MWT | 10 | 376.9 (87.6) | 36 | 266.8 (111) | -110.1 (-187.2 – 32.9) | <0.001 |
| WHODAS | 59 | 2 (1 – 4) | 109 | 9 (4 – 18) | 7 (4 – 9) | <0.001 |
| SEE | 59 | 72 (61 – 77) | 109 | 54 (44 – 67) | -14 (-19 – 9) | <0.001 |
| EQ-5D-5L | 59 | 0.892 (0.801 – 0.942) | 109 | 0.733 (0.634 – 0.850) | - 0.159 (-0.185 - - 0.088) | <0.001 |

**Supplementary Table 4**. Outcome comparison between prehab ‘Universal’ vs ‘Targeted’ arms

| Variable^α^ | Universal | | Targeted | |  |  |
| --- | --- | --- | --- | --- | --- | --- |
|  | n | Average Change | n | Average Change | Difference  (95% CI) | P-value |
| STS (60s) | 22 | 4.1 (2.1) | 29 | 2.6 (1.9) | 1.494 (-2.630 - -0.3568) | 0.0111 |
| 6MWT | 6 | 31 (34) | 19 | 30 (22) | -1.14 (-25.6 – 23.3) | 0.9241 |
| WHODAS | 42 | 0 (-1 – 0) | 88 | -2 (-4 – 0) | -2 (-2 – 0) | 0.0003 |
| SEE | 42 | 9 (0 – 20) | 85 | 9 (0 – 16.5) | 0 (-5 – 3) | 0.7535 |
| EQ-5D-5L | 42 | 0 (0 – 0.0675) | 88 | 0.001 ( 0 - 0.052) | 0.001 ( -0.0120 – 0.005) | 0.9684 |

**Supplementary Table 5:** Grade 2 Toxicity Assessment Regression (univariate only)

| **Characteristic** | **N** | **OR** **(95% CI)***^1^* | **p-value** |
| --- | --- | --- | --- |
| Age | 318 | 0.97 (0.94 to 0.99) | 0.019 |
| Male | 318 | 0.74 (0.47 to 1.15) | 0.18 |
| IMD | 318 | 1.00 (0.98 to 1.01) | 0.43 |
| CompletedPrehab | 318 | 0.86 (0.55 to 1.34) | 0.50 |
| PerfStat | 318 | 0.95 (0.72 to 1.25) | 0.72 |
| StagesAmmend | 318 | 3.27 (2.47 to 4.40) | <0.001 |
| SACT | 318 | 6.92 (3.64 to 14.2) | <0.001 |
| Fractions | 318 | 1.13 (1.10 to 1.17) | <0.001 |
| *^1^* OR = Odds Ratio, CI = Confidence Interval | | | |

**Supplementary Table 6** - 90 Day Mortality (univariate only)

| **Characteristic** | **N** | **OR** **(95% CI)***^1^* | **p-value** |
| --- | --- | --- | --- |
| **Age** | 318 | 1.06 (0.99 to 1.15) | 0.12 |
| **Male** | 318 | 2.03 (0.62 to 7.72) | 0.26 |
| **IMD** | 318 | 0.97 (0.93 to 1.00) | 0.084 |
| **CompletedPrehab** | 318 | 0.70 (0.20 to 2.23) | 0.54 |
| **PerfStat** | 318 | 1.65 (0.80 to 3.52) | 0.18 |
| **StagesAmmend** | 318 | 1.65 (0.87 to 3.33) | 0.13 |
| **SACT** | 318 | 1.26 (0.27 to 4.37) | 0.73 |
| **Fractions** | 318 | 0.99 (0.93 to 1.06) | 0.85 |
| *^1^* OR = Odds Ratio, CI = Confidence Interval | | | |

**Supplementary Table 7** - 365 Day Mortality (univariate and multivariate)

| **Characteristic** | **Univariable** | | | **Multivariable** | |
| --- | --- | --- | --- | --- | --- |
|  | **N** | **OR** **(95% CI)***^1^* | **p-value** | **OR** **(95% CI)***^1^* | **p-value** |
| Age | 318 | 1.03 (0.99 to 1.07) | 0.18 | 1.04 (1.00 to 1.09) | 0.048 |
| Male | 318 | 2.13 (1.13 to 4.17) | 0.022 | 2.60 (1.30 to 5.42) | 0.008 |
| IMD | 318 | 1.00 (0.98 to 1.01) | 0.69 |  |  |
| CompletedPrehab | 318 | 0.51 (0.26 to 0.95) | 0.038 | 0.50 (0.24 to 1.00) | 0.054 |
| PerfStat | 318 | 1.91 (1.28 to 2.91) | 0.002 | 2.67 (1.69 to 4.33) | <0.001 |
| StagesAmmend | 318 | 1.97 (1.39 to 2.86) | <0.001 | 2.86 (1.88 to 4.50) | <0.001 |
| SACT | 318 | 1.35 (0.63 to 2.70) | 0.42 |  |  |
| Fractions | 318 | 1.02 (0.98 to 1.05) | 0.29 |  |  |
| *^1^* OR = Odds Ratio, CI = Confidence Interval | | | | | |

**Supplementary Table 8.** Differences of outcome by lockdown status.

| Variable^α^ | During COVID lockdown | | End of Lockdown | |  |  |
| --- | --- | --- | --- | --- | --- | --- |
|  | n | Average^β^ | n | Average^β^ | Difference  (95% CI) ^δ^ | P-value^δ^ |
| STS (60s) | 11 | 2.09 (2.16) | 40 | 3.6 (1.97) | 1.51 (0.13 – 2.86) | 0.0324 |
| WHODAS | 39 | -1 (-4 – 0) | 93 | -1 (-2.5 – 0) | 0 (-1 – 1) | 0.8778 |
| SEE | 39 | 9 (2 – 15) | 93 | 9 (0 – 18) | 0 (-5 – 3) | 0.5805 |
| EQ-5D-5L | 39 | 0.013 (0 – 0.063) | 93 | 0 (0 – 0.058) | 0 (-0.021 – 0) | 0.3268 |

α; STS, Sit-to-Stand test; WHODAS, WHO Disability Assessment Schedule 2.0; SEE, Self-Efficacy for Exercise Scale; IPAQ, International Physical Activity Questionnaire; EQ-5D-5L, EuroQul-5 Dimension-5 Level assessment;
β Mean (standard deviation); Median (Interquartile Range)

δPaired values assessed, Paired T-Test (parametric data), Wilcoxon Signed Rank (non-parametric data)
